# A Data-Driven Approach to Polycystic Ovary Syndrome Diagnosis: Evaluating Machine Learning Models

**DOI:** 10.1101/2025.07.13.25331465

**Authors:** Payam Mohammadi, Najmeh Parvaz, Mohammad Masoud Eslam, Sara Zareei

## Abstract

**Background:** PCOS is recognized as a major health concern affecting women around the world. Early detection and treatment of PCOS significantly reduce implications in the future. Conventional diagnostic methods are resource-intensive and may be prone to inaccuracies. We should utilize early diagnostic techniques to reduce the severity and overall impact. Machine learning offers a promising approach to improving PCOS detection by analyzing clinical and demographic data efficiently.

**Methods:** This study utilized a dataset of 539 women, including 176 PCOS-positive cases, sourced from the Kaggle repository. Thirty-eight features, categorized into anthropometric, symptom-based, test result, and demographic variables, were analyzed. The most important Feature importance was assessed using the Mean Squared Error metric. Six machine learning models were employed to classify PCOS cases.

**Results:** Significant differences were observed in multiple clinical and anthropometric variables between PCOS-positive and PCOS-negative cases, including BMI, waist-to-hip ratio, antral follicle count, AMH levels, and menstrual cycle length. The most predictive features were antral follicle count, hair growth, skin pigmentation, weight gain, and fast-food consumption. Among all models, Random Forest, the highest-performing model, demonstrated the efficacy of machine learning in PCOS prediction with a 93% accuracy and 86% high sensitivity.

**Conclusions:** Machine learning can improve early and accurate PCOS detection, providing a cost-effective and efficient substitute for traditional methods of diagnosis. The integration of predictive models into clinical practice could facilitate timely interventions, improving patient outcomes and reducing the healthcare burden associated with PCOS.

## Introduction

Polycystic ovary syndrome (PCOS) is one of the most common endocrine disorders, impacting approximately 5% to 10% of women of childbearing age (1). This syndrome is associated with an increased risk of endometrial hyperplasia and endometrial cancer, cardiovascular disease, diabetes, infertility, and metabolic syndrome (2). Symptoms of PCOS may vary among individuals, but common manifestations include irregular menstrual cycles, hirsutism, acne, hormonal imbalances (elevated male hormone levels), and ovarian structural changes. Additionally, some individuals may experience weight gain, infertility, skin blemishes, and significant hair loss (3).

The diagnosis of this condition involves a comprehensive evaluation, including a review of medical history, physical examination, blood tests, ultrasound imaging, and differential diagnosis to rule out other causes. While thorough, this process can be resource-intensive and prone to human error, which may affect accuracy. A precise diagnosis is crucial for developing effective, personalized treatment plans, ultimately improving clinical outcomes (4).

Predictive medicine is a rapidly evolving field that leverages advanced technologies, such as machine learning, artificial intelligence, and genomics, to anticipate the likelihood of diseases before they manifest (5-9). By analyzing vast amounts of patient data, including genetic information, biomarkers, lifestyle factors, and medical history, predictive models can identify individuals at risk for conditions like cancer, cardiovascular diseases, and neurodegenerative disorders (10-13). This proactive approach enables early intervention, personalized treatment plans, and preventive strategies, ultimately improving patient outcomes and reducing healthcare costs (14-18). As precision medicine continues to advance, predictive medicine is poised to play a crucial role in transforming healthcare by shifting the focus from reactive treatments to proactive disease prevention (19-22).

Machine learning (ML) has become indispensable in contemporary healthcare environments, facilitating the analysis of extensive datasets generated in clinical settings (23). These approaches enhance diagnostic capabilities by effectively processing diverse and large-scale medical data (24). Through the utilization of machine learning algorithms, PCOS can be detected earlier, before it advances to a severe state. Various learning algorithms, such as the K-nearest neighbor (KNN), Support Vector Machine (SVM), Logistic Regression (LR), Decision Tree (DT), Random Forest (RF), Linear Discriminant Analysis (LDA), AdaBoost (AB), XGBoost (XB), and CatBoost algorithms are used to predict PCOS (25-27).

This investigation aims to explore the application of a variety of machine learning methodologies, including LR, SVM, NB, RF, KNN, and DT, for the identification of a particular health condition. The study seeks to contribute to the advancement of detection methods by providing more accurate and reliable tools, which can enhance the quality of life for individuals affected by this syndrome.

## Methods

### Study design

The present study included data from 541 participants gathered in ten hospitals across Kerala, India. After excluding inconsistent participants, a total of 539 individuals were included in the study, of whom 176 were identified as PCOS-positive cases. Health-related information for each participant was collected and categorized into anthropometric, symptomatic, test result, and demographic variables.

### Anthropomorphic variables

In the present study, the four anthropometric variables are hip circumference, waist circumference, waist-to-hip ratio, and body mass index (BMI).

### Symptom variables

Symptom variables consist of six variables: cycle length, weight gain, unwanted hair growth, Skin darkening, hair loss, and pimples.

### Test result variables

The test result variables include pulse rate, respiratory rate (RR), hemoglobin (HB) level, human chorionic gonadotropin I (IbetaHCG), human chorionic gonadotropin II (IIbetaHCG), follicle-stimulating hormone (FSH), luteinizing hormone (LH), the FSH to LH ratio (FSH/LH), prolactin (PRL), vitamin D3 (Vit D3), random blood sugar (RBS), progesterone (PRG), anti-Müllerian hormone (AMH), thyroid-stimulating hormone (TSH), systolic and diastolic blood pressure (BP), blood group (BG), the number of antral follicles in the right and left ovaries, average follicle size in the right and left ovaries, and endometrial thickness.

### Demographic variables

The variables, including age, marital status, abortion history, pregnancy status, regular exercise activity, and fast-food consumption, are classified as demographic variables.

### Statistical and machine learning approaches

Machine learning models are applied to classify individuals into two groups based on PCOS status: PCOS-negative and PCOS-positive cases. Various machine learning algorithms, including LR, SVM, NB, RF, KNN, and DT, are employed to predict PCOS.

LR is a classification algorithm used to estimate binary outcomes based on independent variables, predicting the probability of events by fitting data to a logit function, with outcome values between 0 and 1.

The SVM algorithm represents data points as vectors in an n-dimensional space, with support vectors located near the margin of the classifier (28).

In Naïve Bayes, features are assumed to be independent.

KNN is a non-parametric model that predicts outcomes based on similar data points and is used for various tasks such as classification, regression, text categorization, image recognition, and clustering, utilizing three types of distance metrics (29).

DT is also a non-parametric algorithm where nodes represent input attributes, internal nodes indicate factors, branches show outcomes, and leaf nodes determine classes (30).

The Random Forest model consists of a group of decision trees that collectively make predictions. Each tree votes for the class based on attributes.

The dataset, comprising 38 features, was randomly split into 80% training data and 20% test data. A 5-fold cross-validation approach was applied for internal validation. Model performance was assessed using six metrics: sensitivity, specificity, accuracy, positive predictive value (PPV), and negative predictive value (NPV) (31-33). Feature importance was evaluated based on Mean Squared Error (MSE) (34, 35). R software and T-tests were used for statistical analysis, with a p-value < 0.05 considered statistically significant.

### Data Availability

The data used for the purpose of this study were available in Kaggle website: https://www.kaggle.com/datasets/prasoonkottarathil/polycystic-ovary-syndrome-pcos

## Results

### Study characteristics

In this research, data from 539 women were extracted from the Kaggle dataset. Among the collected data, 176 cases were PCOS-positive. The average age of PCOS-negative cases was 32.08 years, while the average age of PCOS-positive cases was 30.09 years. Highly significant differences (P < 0.001) were found across all six symptom variables, BMI, hip and waist measurements, antral follicle count in both ovaries, AMH levels, age, and fast-food intake. Additionally, significant differences were observed in average follicle size between the groups (Table 1).

**Table 1.** Comparison between the PCOS -positive group and the PCOS-negative group with P values (N=539).

| Characteristics | PCOS-Negative Cases<br>(n=363) | PCOS-Positive Cases<br>(n=176) | P-value |
| --- | --- | --- | --- |
| Age (mean (SD)) | 32.08 (5.36) | 30.09 (5.29) | <0.001 |
| BMI (mean (SD)) | 23.75 (3.76) | 25.47 (4.41) | <0.001 |
| Pulserate (mean (SD)) | 72.97 (5.04) | 73.84 (2.74) | 0.032 |
| Hb (mean (SD)) | 11.11 (0.88) | 11.27 (0.83) | 0.044 |
| Cycle length (mean (SD)) | 5.13 (1.26) | 4.55 (1.82) | <0.001 |
| Marriage Status (mean (SD)) | 8.07 (4.82) | 6.90 (4.70) | 0.008 |
| Hip (mean (SD)) | 37.55 (3.78) | 38.91 (4.20) | <0.001 |
| Waist (mean (SD)) | 33.43 (3.45) | 34.68 (3.76) | <0.001 |
| AMH (mean (SD)) | 4.54 (4.30) | 7.85 (7.81) | <0.001 |
| VitD3 (mean (SD)) | 29.31 (12.39) | 92.73 (605.64) | 0.046 |
| Weight gain (%) | 83 (22.9) | 121 (68.8) | <0.001 |
| Unwanted hair growth (%) | 47 (12.9) | 101 (57.4) | <0.001 |
| Skindarkening (%) | 55 (15.2) | 110 (62.5) | <0.001 |
| Hair loss (%) | 142 (39.1) | 102 (58.0) | <0.001 |
| Pimples (%) | 142 (39.1) | 123 (69.9) | <0.001 |
| Fast food (%) | 139 (38.3) | 139 (79.0) | <0.001 |
| nFollicleL (mean (SD)) | 4.35 (2.82) | 9.76 (4.31) | <0.001 |
| nFollicleR (mean (SD)) | 4.63 (2.93) | 10.80 (4.15) | <0.001 |
| AvgFollicleL (mean (SD)) | 14.70 (3.87) | 15.68 (2.73) | 0.003 |
| AvgFollicleR (mean (SD)) | 15.23 (3.41) | 15.90 (3.09) | 0.029 |
| Endometrium (mean (SD)) | 8.31 (2.26) | 8.81 (1.93) | 0.011 |

### Classification performance

The dataset, incorporating 38 features, was split into 80% training data and 20% test. Six ML models including LR, SVM, NB, RF, KNN, and DT were applied to these features. The results of analysis are demonstrated in Figure1. Most of the models provided progressive results according to sensitivity, specificity, accuracy, positive predictive values, and negative predictive values. In terms of accuracy, Random Forest and Logistic Regression achieved success rates of more than 90% (93% and 92%, respectively). All models showed specificity greater than 90%, with the Naïve Bayes and Logistic

**Figure 1.**
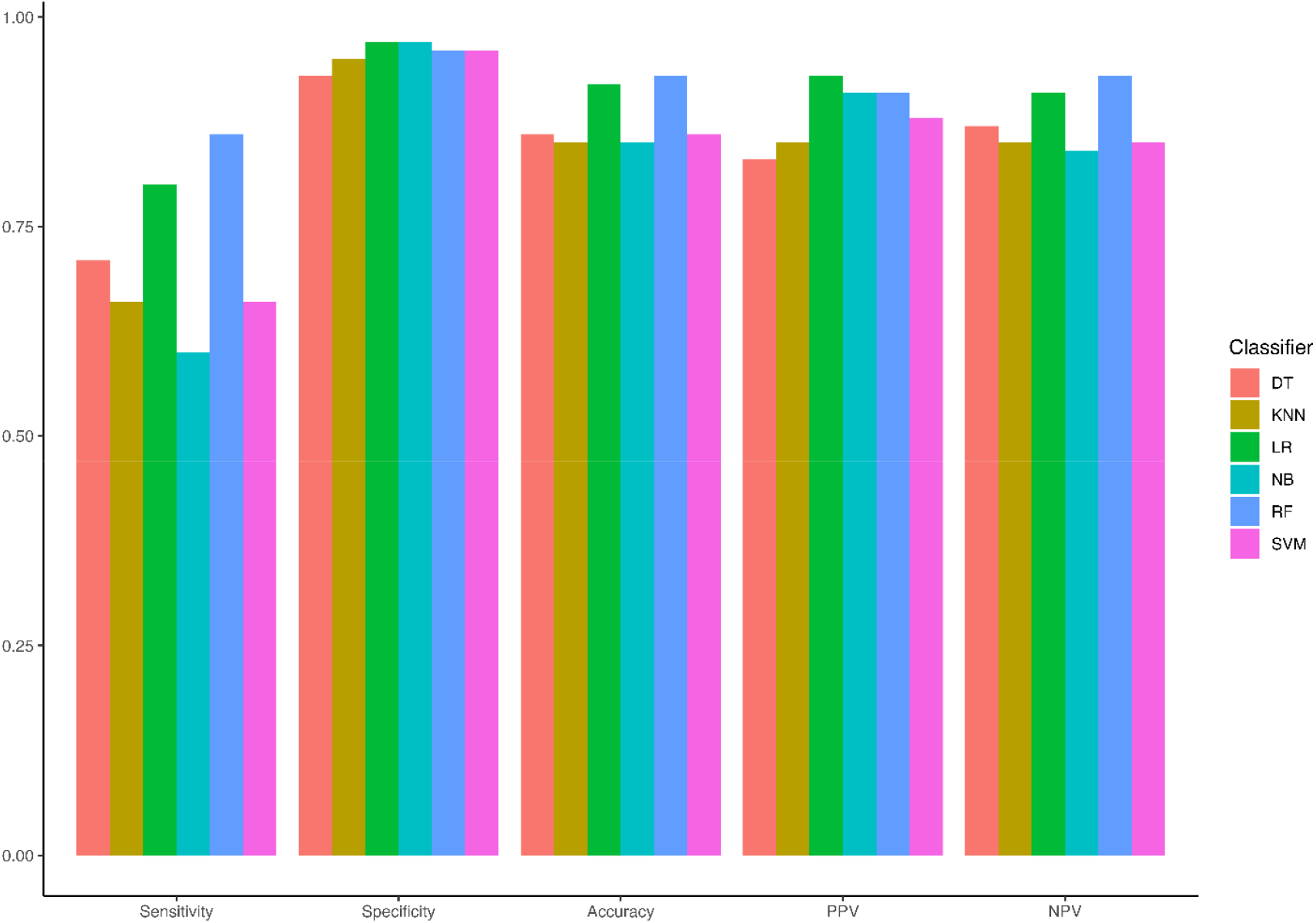
Comparative performance of machine learning models based on sensitivity, specificity, accuracy, PPV, and NPV for each model

Regression models having the highest specificity (97%). Among all models, Random Forest had the highest sensitivity (86%), while Naïve Bayes showed the lowest sensitivity (60%). Models including Naïve Bayes, Random Forest, and Logistic Regression exhibited PPV greater than 90%. The highest NPV values were observed for Random Forest and Logistic Regression, with values of 93% and 91%, respectively.

### Features importance analysis

To determine the most important features, the Mean Squared Error (MSE) metric was used. In Figure 2, the most important features, ranked according to their importance level, are presented. The most influential features included the number of antral follicles in the right and left ovaries, hair growth, skin darkening, weight gain, fast food consumption, cycle length, hip circumference, AMH levels, and pimples, respectively (Figure 2). These features were found to have the highest impact on model predictions, demonstrating their association with PCOS prediction. The number of antral follicles in the right ovary, with an importance value of 43.3%, was identified as the most significant feature compared to other factors in the prediction of this disease.

**Figure 2.**
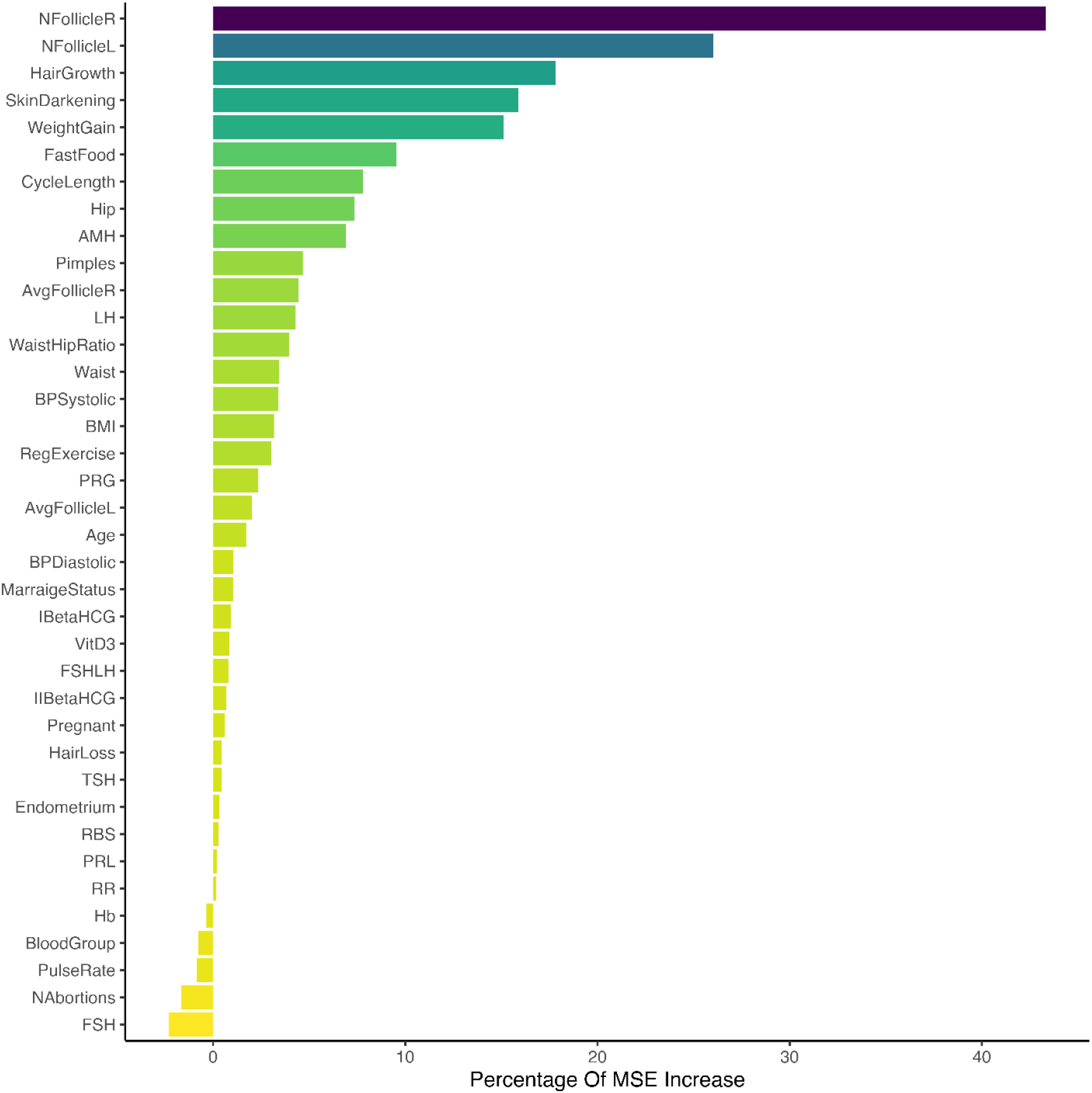
Feature importance for all variables according to the MSE metric

## Discussion

Polycystic Ovary Syndrome is a prevalent and persistent hormonal condition affecting females. The complexity of the syndrome renders its identification challenging. Moreover, deficiencies in early identification impede women from effectively managing this chronic condition. Therefore, computer-aided diagnostic devices are crucial for facilitating earlier diagnoses (36). Machine learning techniques have become essential tools for analyzing the vast amounts of healthcare data generated in modern medical settings. These techniques enable precise disorder diagnosis by effectively evaluating clinical datasets (37). This study is designed to examine the utilization of diverse machine learning techniques, including LR, SVM, NB, RF, KNN, and DT, for the detection of PCOS based on the health measures of 539 female participants.

In this research, anthropometric variables, including BMI, hip and waist measurements, all symptom variables, AMH levels, antral follicle count in both ovaries, age, and fast-food intake, were significantly different between the two groups. These findings align with prior studies that reported anthropometric variables (38), symptom variables, AMH levels (39), antral follicle count in both ovaries, age, and dietary habits as important factors in the diagnosis of PCOS.

The application of six machine learning models to 38 features demonstrated that the Random Forest model is a highly effective method for predicting PCOS in this study, achieving an accuracy of 93%. According to the present results, one study revealed that among NB, KNN, LR, RF, and SVM models, the most accurate method for the prediction of PCOS was RF, achieving an accuracy of 89.02% (40). In another study, the authors compared various classifiers, including RF, KNN, SVM, and DT, for PCOS identification. The Random Forest model exhibited the highest performance, with an accuracy of 93.25% (41). The hybrid Random Forest and Logistic Regression (RFLR) model achieved the highest accuracy of 91.01%, utilizing 5-fold cross-validation on the 10 most significant features to predict PCOS (42). In research involving 541 women, RF was found to be the best model for predicting PCOS, achieving an accuracy of 86% compared to other machine learning algorithms (43). Additionally, it has been reported that the RF model outperformed other algorithms, including KNN, SVM, and NB classifiers, with an accuracy of 93.5% (25). In one study, the researchers employed various machine learning techniques to identify patients with PCOS. Analysis of the results revealed that the Random Forest algorithm demonstrated exceptional performance, achieving 96% accuracy in diagnosing PCOS on the evaluated dataset (44). Another study employed SVM, LR, NB, and KNN to detect PCOS in women. The chi-square feature selection method was used to identify the top 30 features, with Random Forest achieving the highest accuracy (45). These findings demonstrate the efficacy of the RF algoritm in accurate PCOS prediction.

Although the results of this study are consistent with studies reporting RF as an effective algorithm for PCOS prediction, they differ from some published studies that identified CatBoost Classifier (46), DT (47), and SVM (48) as achieving the highest accuracy in predicting PCOS. Possible explanations for variations in machine learning results for PCOS prediction include differences in datasets, feature selection methods, preprocessing techniques, validation strategies, and performance metrics.

The Mean Squared Error metric was utilized to identify the most influential factors in predicting PCOS. The findings reveal that the number of antral follicles in both the right and left ovaries, hair growth, skin pigmentation, weight gain, fast food consumption, menstrual cycle length, hip circumference, AMH levels, and acne were among the most significant predictors. The present findings are consistent with previous works identifying antral follicle count as a critical diagnostic marker for PCOS due to its strong association with ovarian dysfunction and increased follicular reserve (49). This reinforces the assertion that ovarian morphology, particularly an increased follicle count, serves as a key diagnostic indicator of PCOS, as defined by the Rotterdam criteria (50). Elevated level of AMH is a key indicator of PCOS and has been identified as a significant feature in the present study. Previous research has reported that an increase in antral follicle count is associated with elevated AMH levels (49). Moreover, higher serum AMH levels are associated with ovulatory dysfunction and hyperandrogenism, indicating that AMH may impair follicular development in PCOS (51).

A significant prevalence of dermatological symptoms, including hirsutism and skin pigmentation, has been documented in patients with PCOS. Insulin resistance and hyperandrogenism play a crucial role in the development of these symptoms (52). Insulin resistance, weight gain, and hormonal imbalances that impede reproductive function can come from poor diets, especially fast food (53). Variations in cycle duration indicate menstrual irregularities, a key indicator of PCOS ovulatory dysfunction (54).

In conclusion, PCOS is a prevalent and complex hormonal condition that affects women’s reproductive health. The diverse symptoms and presentations of PCOS make it challenging to identify and manage. However, machine learning techniques, particularly Random Forest, have shown promise in improving the accuracy of early diagnosis, leading to cost reductions and a diminished burden of PCOS on both patients and the healthcare system.

## Data Availability

All data produced in the present study are available upon reasonable request to the authors

## Acknowledgements

NA

